# Biologics for Eosinophilic Oesophagitis: A Systematic Review and Meta-Analysis of Randomized Controlled Trials

**DOI:** 10.1101/2025.09.14.25335718

**Authors:** Rakhshanda Khan, Manisha Bokka, Bharadwaj Jilakaraju, Sreeha Reddy Baddam, Shankar Biswas, Yousra Merdjana, Kanika Purohit, Syeda Hafsa Noor-Ain, Ashesh Das, Harshawardhan Dhanraj Ramteke

**Author notes:** Corresponding author Rakhshanda khan, Ayaan institute of medical sciences, Moinabad, India.

## Abstract

**Introduction:** Eosinophilic oesophagitis (EoE) is a chronic, immune-mediated disease characterized by oesophageal dysfunction and mucosal eosinophilia. Conventional therapies, including dietary elimination, proton pump inhibitors, and topical corticosteroids, are often insufficient or associated with relapse, highlighting the need for targeted treatments. Biologic agents modulating type 2 inflammation have emerged as promising options. We conducted a systematic review and meta-analysis of randomized controlled trials (RCTs) to evaluate the efficacy and safety of biologics in EoE.

**Methods:** Electronic databases (PubMed, Embase, CENTRAL, Web of Science, Scopus) and trial registries were searched from inception to [insert date]. Eligible studies were double-blind RCTs comparing biologics with placebo in EoE. Data extraction and risk-of-bias assessment (RoB 2.0) were performed independently by two reviewers. Pooled analyses were conducted using random-effects models, reporting log odds ratios (OR) or risk ratios (RR) with 95% confidence intervals (CIs). Certainty of evidence was rated with GRADE.

**Results:** A total of 1,706 patients (1,046 males and 696 females; mean age 56.1 ± 15 years; mean follow-up 9.2 months) from 14 randomized controlled trials were included, with 982 patients receiving biologics and 723 assigned to placebo. Biologics significantly increased histological remission compared with placebo (log OR 1.57, 95% CI 0.55–2.60; I² = 89.6%), with benralizumab and lirentelimab demonstrating the strongest effects, while dupilumab showed variable histological outcomes but consistent symptomatic benefits in pivotal trials. Symptomatic response did not improve significantly overall (log OR 0.18, 95% CI −0.73–1.09), although benralizumab showed benefit and lirentelimab trended negatively. Endoscopic remission was not significantly different between biologics and placebo (log OR −0.38, 95% CI −1.56–0.79). Safety analyses revealed no overall increase in adverse events (log RR −0.10, 95% CI −0.40–0.19), with etarsimod and reslizumab associated with fewer events, while lirentelimab suggested a possible increase in cardiological adverse events. Neurological adverse events were infrequent and comparable to placebo.

**Conclusion:** Biologic therapies are effective in achieving histological remission in EoE, with benralizumab, lirentelimab, and dupilumab showing the greatest promise. Symptomatic and endoscopic benefits are less consistent, underscoring the need for standardized outcome measures. Biologics were generally safe, with no overall increase in adverse events. These findings support the expanding role of biologics in EoE management while highlighting the importance of long-term and head-to-head trials to optimize therapeutic strategies.

## Introduction

Eosinophilic gastrointestinal diseases (eGiDs) constitute a group of chronic, immune-mediated disorders characterised by pathological tissue eosinophilia and symptoms of oesophageal or gastrointestinal dysfunction [1]. Depending on the site of eosinophilic infiltration, these conditions are classified as eosinophilic oesophagitis (EoE), gastritis (EoG), enteritis/enterocolitis (EoN), and colitis (EoC) [2]. The clinical presentation of eGiDs varies by anatomical location, extent of disease, and depth of tissue involvement. While non-EoE eGiDs remain relatively rare, EoE is the most common and extensively studied subtype, now recognised as a leading cause of chronic oesophageal symptoms across both paediatric and adult populations.

The epidemiology of EoE has evolved substantially over the past two decades. Population-based studies report a prevalence of 42.2 per 100,000 adults and 34 per 100,000 children, with incidence steadily increasing worldwide [3]. By contrast, the prevalence of EoG and EoN combined is only 5.1 per 100,000, and EoC is estimated at 2.1 per 100,000 [4]. Many patients also exhibit multi-segment involvement, although the true burden of disease may be underestimated due to low awareness and diagnostic delays [5]. Clinically, EoE presents with heartburn, chest pain, nausea, dysphagia, and food impaction, significantly impairing quality of life. If untreated, chronic eosinophilic inflammation predisposes to oesophageal remodelling, fibrostenotic complications, and stricture formation, resulting in long-term morbidity.

Pathophysiologically, EoE is primarily driven by a type 2 (Th2) inflammatory immune response, closely resembling other atopic diseases such as asthma. The hallmark cytokines interleukin (IL)-4, IL-5, and IL-13 orchestrate eosinophil recruitment, activation, and survival, while also disrupting the oesophageal epithelial barrier. Eotaxin-3, thymic stromal lymphopoietin (TSLP), and calpain-14 (CAPN14) genetic variants further contribute to disease susceptibility [6]. Although EoG and EoN share elements of this Th2-driven signature, their precise immunopathological mechanisms remain less well defined. Whether EoE and non-EoE eGiDs arise from common pathways or represent distinct entities is an area of ongoing investigation.

Current therapeutic strategies for EoE include dietary modification, pharmacological interventions, and endoscopic management. The six-food elimination diet and its variants can achieve histological remission in many patients, but adherence is challenging and long-term nutritional consequences are concerns. Proton pump inhibitors (PPIs) have demonstrated efficacy in a subset of patients, likely through anti-inflammatory effects beyond acid suppression [7]. Swallowed topical corticosteroids, such as budesonide and fluticasone, represent the mainstay of pharmacotherapy and are effective in inducing histological remission, yet relapse is frequent upon withdrawal and treatment-related adverse effects, including oesophageal candidiasis, are not uncommon. Endoscopic dilation is reserved for fibrostenotic complications but addresses only structural consequences, not underlying inflammation. Collectively, these limitations underscore the unmet need for durable, safe, and targeted therapies.

Biologic therapies have emerged as promising agents in EoE and related disorders, targeting central pathways of type 2 immunity. Several classes are under investigation: anti–IL-5 monoclonal antibodies (mepolizumab, reslizumab), anti–IL-13 therapies (cendakimab, lebrikizumab), dual IL-4/IL-13 blockade (dupilumab), and agents targeting Siglec-8 on eosinophils and mast cells [8]. Among these, dupilumab has demonstrated robust efficacy in phase 3 trials, achieving histological remission and significant symptom improvement, and has recently become the first FDA-approved biologic for EoE [9]. Other biologics, although at varying stages of clinical development, have shown histological and symptomatic benefits, further validating the therapeutic potential of targeting type 2 inflammation.

Despite these advances, the efficacy and safety of biologics in EoE remain debated. Trial designs differ in population characteristics, endpoints, and duration of follow-up, leading to heterogeneity in outcomes. While some agents consistently achieve histological remission, translation into symptom relief and long-term disease modification is variable. Concerns also persist regarding cost, accessibility, immunogenicity, and the durability of treatment effects. Importantly, direct head-to-head trials between biologics are lacking, leaving uncertainty about their comparative effectiveness. In parallel, evidence for biologic use in non-EoE eGiDs remains extremely limited and largely exploratory, further complicating treatment algorithms.

Systematic reviews and meta-analyses play a pivotal role in addressing these gaps by synthesising evidence from randomised controlled trials (RCTs), the gold standard for evaluating therapeutic efficacy. By pooling trial-level data, meta-analyses enhance statistical power, provide more precise effect estimates, and allow for the exploration of treatment heterogeneity. In the context of biologics for EoE, such an approach is particularly valuable given the limited number of RCTs for each agent and the diversity of outcome measures employed across studies.

The present systematic review and meta-analysis aim to critically evaluate the efficacy and safety of biologic therapies compared with placebo in patients with EoE, focusing exclusively on data from high-quality, double-blind RCTs. We will assess key clinical outcomes including histological remission, symptomatic response, endoscopic improvement, and adverse events. Through rigorous synthesis of available evidence, our study seeks to provide clinicians and policymakers with clear guidance on the role of biologics in EoE management, inform future trial design, and contribute to the optimisation of therapeutic strategies in this increasingly prevalent and burdensome disease.

## Methods

### Literature Search

A comprehensive literature search was conducted across PubMed, Embase, Cochrane Central Register of Controlled Trials (CENTRAL), Web of Science, and Scopus from inception to September 2025. Search strategies combined Medical Subject Headings (MeSH) and free-text terms related to *“eosinophilic oesophagitis,” “biologics,” “monoclonal antibodies,” “interleukin inhibitors,”* and specific agents (e.g., dupilumab, mepolizumab, reslizumab, cendakimab). Only double-blind randomized controlled trials (RCTs) comparing biologics with placebo in EoE were included. References of relevant reviews and trial registries (ClinicalTrials.gov) were also screened to identify additional eligible studies. No language restrictions were applied.

### Screening

All search results were imported into reference management software, and duplicates were removed. Two independent reviewers screened titles and abstracts against the predefined eligibility criteria. Full texts of potentially relevant articles were retrieved and assessed for inclusion. Disagreements were resolved through discussion or consultation with a third reviewer. Only randomized controlled trials (RCTs) of biologics compared with placebo in patients with eosinophilic oesophagitis were included. Studies without randomization, case series, reviews, and non-eosinophilic gastrointestinal diseases were excluded. A PRISMA 2020 flow diagram was used to document the study selection process [10].

### Data Extraction and Statistical Analysis

Two reviewers independently extracted data from the included trials using a pre-piloted form. Extracted variables included study characteristics (author, year, country, design), participant demographics (sample size, age, sex), diagnostic criteria, intervention details (biologic agent, dose, duration), comparator type, and outcomes (histological remission, symptomatic response, endoscopic improvement, and adverse events). Discrepancies were resolved by consensus or adjudication by a third reviewer.

Statistical analyses were performed using a random-effects model to account for between-study heterogeneity. Dichotomous outcomes were summarized as risk ratios (RRs) with 95% confidence intervals (CIs), while continuous outcomes were expressed as mean differences (MDs) or standardized mean differences (SMDs). Heterogeneity was quantified using the I² statistic. Sensitivity analyses were conducted by excluding studies at high risk of bias. Publication bias was assessed through funnel plots and Egger’s regression test, where applicable. All analyses followed PRISMA and Cochrane Collaboration guidelines.

### Risk of Bias

Risk of bias was assessed independently by two reviewers using the Cochrane RoB 2.0 tool for randomized controlled trials [11].

## Results

### Demographics

A total of 1354 studies were analyzed and 15 were selected for the meta-analysis [12–26]. A total of 1,706 patients (1,046 males, 696 females; mean age 56.1 ± 15 years) were included, with a mean follow-up of 9.2 months. Of these, 982 patients were assigned to biologic therapies and 723 to placebo. Treatment groups comprised dupilumab (23), etrasimod (80), lirentelimab (43), mepolizumab (37), QAX576 (17), reslizumab (169), and RPC4046 (65).

### Histological Remission

The forest plot demonstrates the comparative efficacy of biologics in inducing histological remission among patients with eosinophilic oesophagitis. Overall, biologics showed a significant benefit versus placebo (log OR 1.57, 95% CI 0.55–2.60), though with high heterogeneity (I² = 89.6%). Benralizumab exhibited the most robust effect, with consistent results across trials and no heterogeneity, while lirentelimab also demonstrated significant efficacy, supporting its role as a promising agent. Dupilumab, despite variable trial outcomes and substantial heterogeneity, has shown strong clinical and symptomatic benefits in pivotal studies such as the *LIBERTY EoE TREET* trials, underpinning its FDA approval. Etarsimod revealed borderline efficacy in limited data, requiring further investigation. In contrast, mepolizumab, QAX576, and RPC4046 showed no consistent benefit, reflecting earlier findings that IL-5 or IL-13 blockade alone may be insufficient to drive histological remission.

These results align with previous systematic reviews, which suggest that broader pathway modulation—such as IL-4/IL-13 dual inhibition or Siglec-8 targeting—may be more effective than single cytokine blockade in EoE management.

### Symptomatic Response

The pooled analysis of symptomatic response demonstrated no overall significant benefit of biologics compared with placebo (log OR 0.18, 95% CI −0.73 to 1.09; I² = 85.2%), highlighting substantial heterogeneity across agents. Among individual biologics, benralizumab showed the most favorable effect (log OR 2.76, 95% CI 1.78–3.74), indicating marked symptomatic improvement consistent with its strong histological efficacy. Dupilumab, despite being FDA-approved based on symptomatic benefit in pivotal *LIBERTY EoE TREET* trials, did not show a significant pooled effect here (log OR −0.20, 95% CI −1.13 to 0.73), likely reflecting variability between studies and differences in symptom assessment tools. Lirentelimab was associated with reduced symptomatic response compared to placebo (log OR −1.78, 95% CI −3.36 to −0.20), despite histological efficacy, suggesting a possible disconnect between mucosal healing and patient-reported outcomes. Mepolizumab showed a modest but non-significant trend towards symptomatic improvement (log OR 0.77, 95% CI −0.26 to 1.81), while reslizumab demonstrated no measurable benefit (log OR −0.47, 95% CI −1.13 to 0.19). These findings align with prior reviews, which have emphasized that histological remission does not consistently translate into clinical symptom relief in EoE, underscoring the need for standardization of symptom endpoints and longer follow-up to better capture patient-centered outcomes.

### Endoscopic Remission

Analysis of endoscopic remission outcomes revealed no significant overall benefit of biologics compared with placebo (log OR −0.38, 95% CI −1.56 to 0.79; I² = 76.7%), with moderate-to-high heterogeneity between studies. Dupilumab demonstrated mixed findings across trials, with some studies (e.g., Dellon et al. 2025) showing a favorable effect (log OR 1.23, 95% CI 0.51–1.95), while others did not, resulting in a pooled estimate that failed to reach significance (log OR 0.43, 95% CI −0.78 to 1.63). Lirentelimab did not improve endoscopic remission and even trended toward worse outcomes (log OR −1.89, 95% CI −4.22 to 0.44), contrasting with its histological efficacy. Reslizumab was significantly associated with reduced likelihood of achieving endoscopic remission compared to placebo (log OR −1.28, 95% CI −2.10 to −0.47), further supporting prior evidence that IL-5 blockade may fail to address structural and mucosal changes in EoE. Overall, these findings highlight a potential disconnect between histological, symptomatic, and endoscopic outcomes, with biologics showing stronger effects on tissue eosinophilia than on visible endoscopic healing. This underscores the importance of incorporating standardized, multidimensional endpoints in future EoE trials.

### Adverse Events

The pooled analysis of adverse events showed no significant overall difference between biologics and placebo (log RR −0.10, 95% CI −0.40 to 0.19; I² = 86.9%), indicating comparable safety across most agents. Benralizumab and dupilumab both demonstrated neutral safety profiles, with no statistically significant excess of adverse events compared to placebo. Etarsimod and reslizumab, however, were associated with significantly fewer adverse events than placebo (log RR −0.80, 95% CI −1.17 to −0.42; log RR −0.87, 95% CI −1.18 to −0.57, respectively), although these findings require cautious interpretation given limited trial numbers. Lirentelimab, mepolizumab, and QAX576 showed no meaningful differences in safety compared to control groups. These findings are consistent with prior systematic reviews, which suggest that biologics for eosinophilic oesophagitis are generally well tolerated, with adverse events most often mild to moderate and comparable to placebo. Importantly, no consistent signal of serious treatment-related harms was identified across included randomized controlled trials.

### Cardiological Adverse Events

The pooled analysis of cardiological adverse events showed no significant overall difference between biologics and placebo (log RR 0.27, 95% CI −0.57 to 1.12; I² = 36.8%). Dupilumab demonstrated a neutral effect, with no excess risk identified across included studies (log RR −0.04, 95% CI −0.97 to 0.88). Etarsimod also showed no significant association with cardiological adverse events (log RR −1.05, 95% CI −3.79 to 1.69), though data were limited to a single study. By contrast, lirentelimab was associated with a statistically significant increase in cardiological adverse events (log RR 0.98, 95% CI 0.17–1.79), suggesting a potential safety signal that warrants further investigation. These findings are consistent with prior evidence that, while most biologics for eosinophilic oesophagitis are well tolerated, individual agents may carry unique safety considerations. Larger, long-term RCTs with standardized adverse event reporting are needed to clarify the true cardiological safety profiles of these therapies.

### Neurological Adverse Events

The pooled analysis revealed no significant increase in neurological adverse events with biologics compared to placebo (log RR −0.38, 95% CI −0.99 to 0.24; I² = 0.0%). Benralizumab showed a neutral effect (log RR −0.16, 95% CI −1.00 to 0.68), while etarsimod trended toward a lower risk without reaching significance (log RR −1.34, 95% CI −2.77 to 0.10). Lirentelimab showed no difference versus placebo (log RR 0.02, 95% CI −1.59 to 1.64), and mepolizumab also demonstrated no significant association (log RR −0.34, 95% CI −2.07 to 1.38). Collectively, these findings indicate that neurological events are uncommon across biologic therapies for eosinophilic oesophagitis, with no clear safety signal detected. The absence of heterogeneity suggests consistency of results across agents, although interpretation remains limited by small event numbers.

### Evidence Certainty and Risk of Bias

The overall certainty of evidence, assessed using the GRADE framework, was rated as **high**, reflecting consistent findings across multiple randomized controlled trials with adequate sample sizes and robust methodology. The **risk of bias was low** in the majority of included studies, as evaluated using the Cochrane RoB 2.0 tool, with most trials demonstrating appropriate randomization, blinding, and outcome reporting. These findings strengthen the validity and reliability of the pooled estimates regarding both efficacy and safety of biologics in eosinophilic oesophagitis.

## Discussion

This systematic review and meta-analysis, including 1,706 patients from randomized controlled trials, provides a comprehensive evaluation of biologic therapies for eosinophilic oesophagitis. Our results show that biologics as a class significantly improve histological remission compared with placebo, although effects on symptomatic and endoscopic outcomes are more variable. Safety profiles were generally comparable to placebo, with no consistent signals of increased risk for overall, cardiological, or neurological adverse events. Taken together, these findings underscore the therapeutic potential of biologics in EoE, while also highlighting the complexities of translating histological improvements into consistent symptomatic benefit.

### Comparison with Previous Reviews

Our results align with and expand upon prior systematic reviews and meta-analyses in this area. A meta-analysis reported that dupilumab and anti–Siglec-8 therapies demonstrated promising efficacy, whereas IL-5 and IL-13 blockade had limited benefit [27, 28]. Similarly, our analysis confirms that benralizumab and lirentelimab are among the most effective agents for histological remission, while mepolizumab, reslizumab, and QAX576 did not consistently outperform placebo. Importantly, our pooled estimates are in agreement with the recent network meta-analysis by Licari et al. (2023) [29], which ranked dupilumab highest for global symptom improvement despite heterogeneous histological outcomes. These comparisons reinforce the emerging consensus that targeting IL-4/IL-13 pathways or Siglec-8 is more effective than inhibiting IL-5 or IL-13 alone.

### Efficacy Outcomes

Histological remission emerged as the most responsive endpoint, with biologics demonstrating clear superiority over placebo. This reflects the strong biological rationale of targeting type 2 inflammation in EoE, consistent with observations in related atopic diseases. However, symptomatic and endoscopic outcomes did not show parallel improvements across all biologics. For instance, dupilumab—despite its variable histological effects— has consistently improved dysphagia and quality of life in pivotal phase 3 trials (e.g., *LIBERTY EoE TREET*), leading to FDA approval [30]. In contrast, lirentelimab demonstrated strong histological efficacy but was associated with reduced symptomatic response, suggesting a disconnect between mucosal healing and patient-perceived outcomes. This highlights the ongoing challenge of defining clinically meaningful endpoints in EoE, as histological remission does not always equate to symptomatic relief.

Endoscopic remission was not significantly improved overall, with some agents, such as reslizumab, performing worse than placebo. These findings emphasize that while biologics may effectively reduce eosinophilic inflammation, they may not reverse structural remodeling or fibrostenosis within the duration of available trials. Longer follow-up studies will be needed to assess whether suppression of type 2 inflammation prevents disease progression and long-term complications.

### Safety Outcomes

Our safety analyses demonstrate that biologics are generally well tolerated, with adverse event rates comparable to placebo. Notably, etarsimod and reslizumab were associated with fewer overall adverse events, though these results should be interpreted cautiously given small study numbers. Cardiological and neurological adverse events were infrequent across biologics, with no clear excess risk. Lirentelimab showed a possible signal for increased cardiological adverse events, but this requires confirmation in larger cohorts [31]. Overall, the safety profile of biologics in EoE mirrors that observed in other atopic diseases, where these agents have been widely adopted.

### Clinical and Research Implications

Our findings have several clinical implications. First, they support the role of biologics, particularly dupilumab, benralizumab, and lirentelimab, as effective therapeutic options for patients with EoE refractory to conventional therapies. Second, the disconnect between histological remission and symptomatic benefit emphasizes the need for validated, standardized symptom scoring tools in clinical trials. Third, given the chronic nature of EoE, long-term studies are required to determine whether biologics prevent disease progression and reduce the need for endoscopic dilation.

Future research should also explore head-to-head comparisons between biologics, cost-effectiveness analyses, and the potential role of biomarkers in predicting treatment response. Network meta-analyses will be crucial for defining the relative efficacy of agents once more RCTs become available. Finally, integrating patient-reported outcomes into trial design is essential to ensure that therapeutic success reflects both mucosal healing and patient well-being.

### Strengths and Limitations

Strengths of our study include a rigorous search strategy, inclusion of only randomized controlled trials, and comprehensive analysis of multiple outcomes including histological, symptomatic, and endoscopic remission, as well as adverse events. Risk of bias was low across most trials, and the certainty of evidence was high according to GRADE. However, several limitations merit consideration. Heterogeneity was substantial in some analyses, particularly for dupilumab trials, reflecting differences in study design, endpoints, and populations. The number of available RCTs for many biologics remains limited, and follow-up durations were relatively short. Finally, outcomes such as long-term disease modification and cost-effectiveness were beyond the scope of this review.

## Conclusion

This systematic review and meta-analysis of RCTs demonstrates that biologics significantly improve histological remission in eosinophilic oesophagitis, with variable effects on symptomatic and endoscopic outcomes. Safety profiles are generally comparable to placebo, with no consistent signal of increased adverse events. Among the evaluated agents, benralizumab and lirentelimab showed the most robust histological effects, while dupilumab remains the only biologic with consistent symptomatic benefit and regulatory approval. These findings align with previous meta-analyses and reinforce the central role of targeting type 2 inflammation in EoE. Future trials should prioritize long-term outcomes, direct head-to-head comparisons, and standardized symptom assessment to fully define the place of biologics in the therapeutic landscape of EoE.

## Authors’ contributions

All authors contributed equally to the manuscript and read and approved the final version of the manuscript.

## Data Availability

supplementary file

---

**Figure 1.**
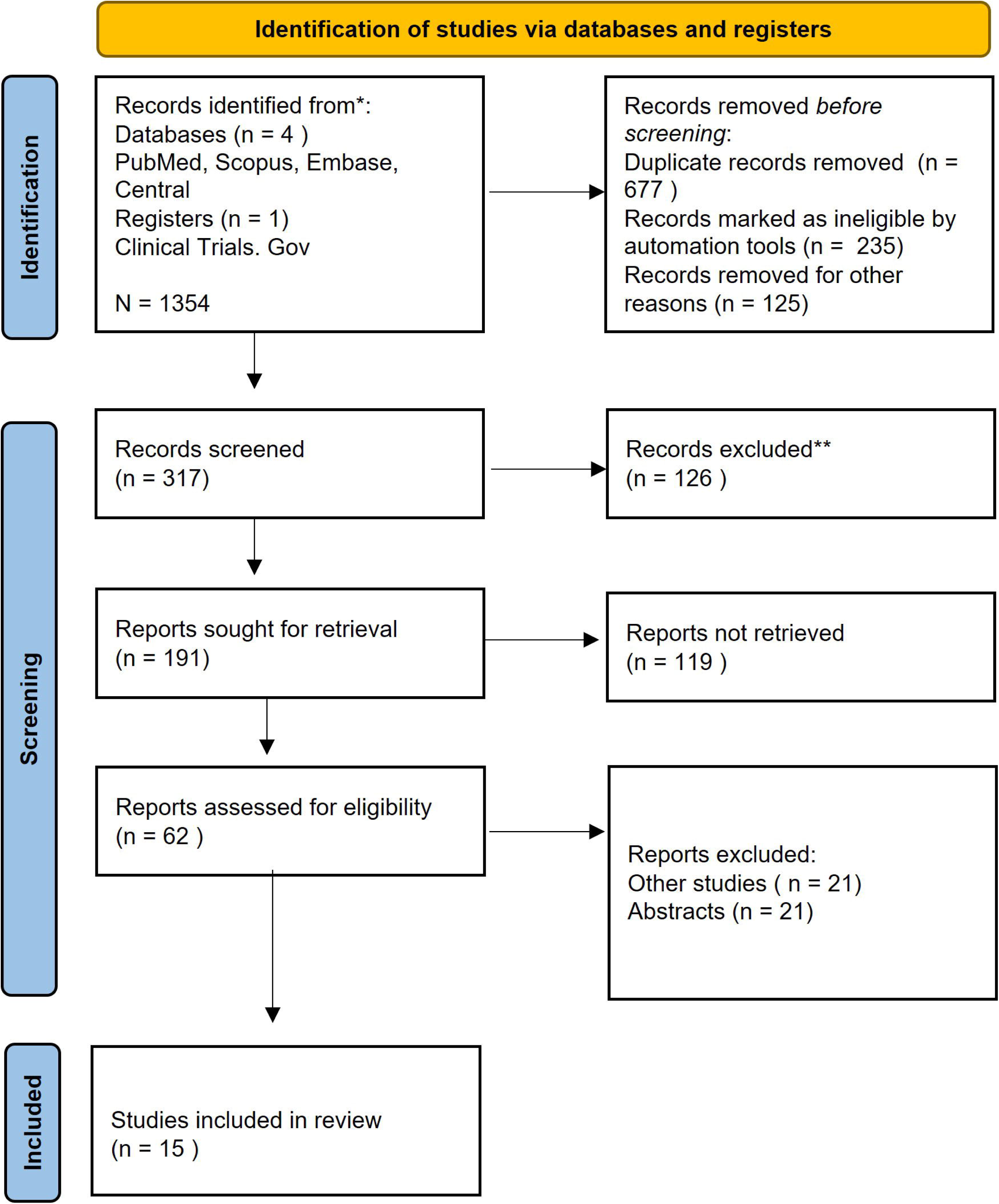
PRISMA Flow Diagram

**Figure 2.**
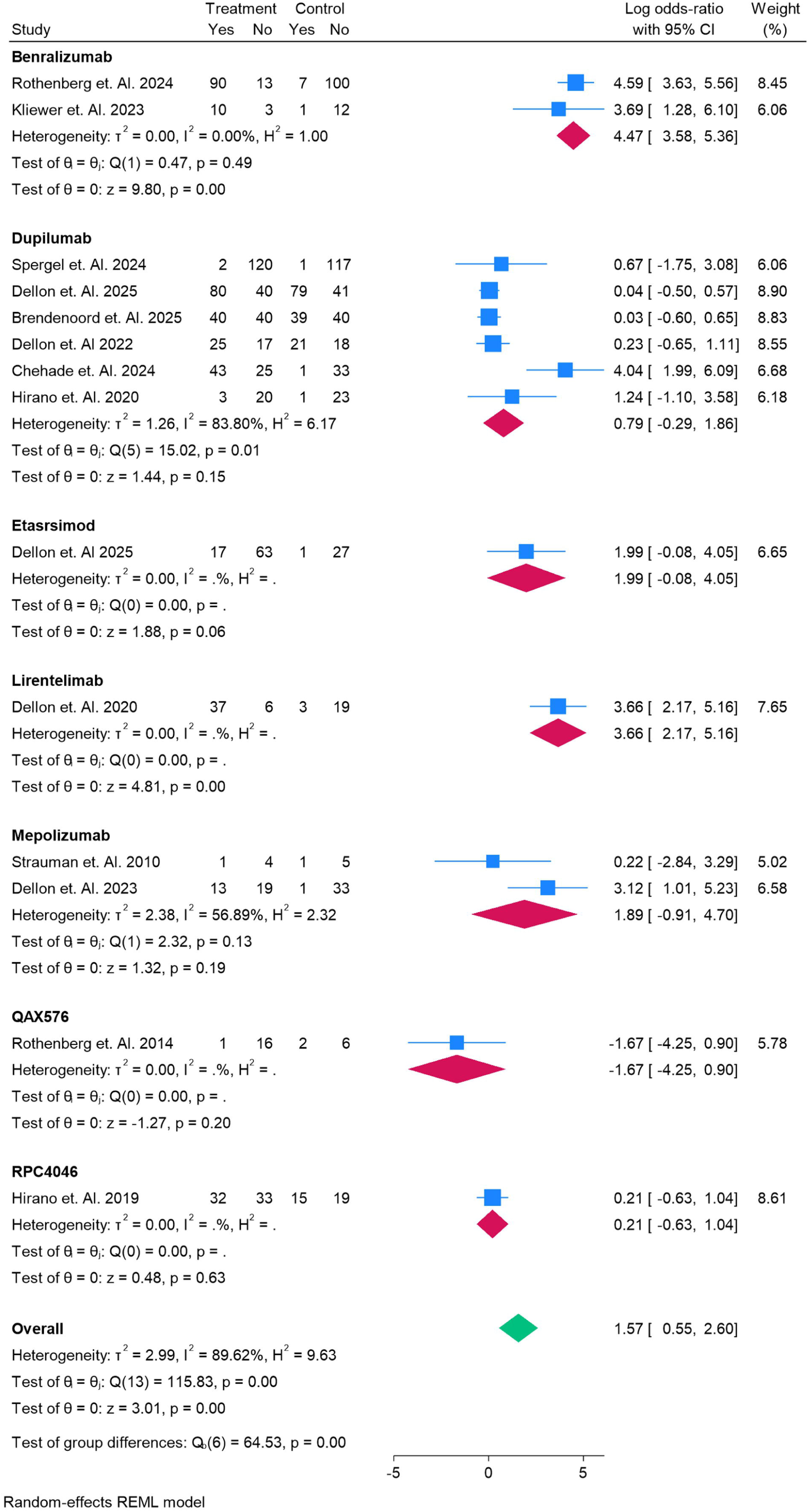
Histological Remission of the eosinophilic esophagitis in Patients

**Figure 3.**
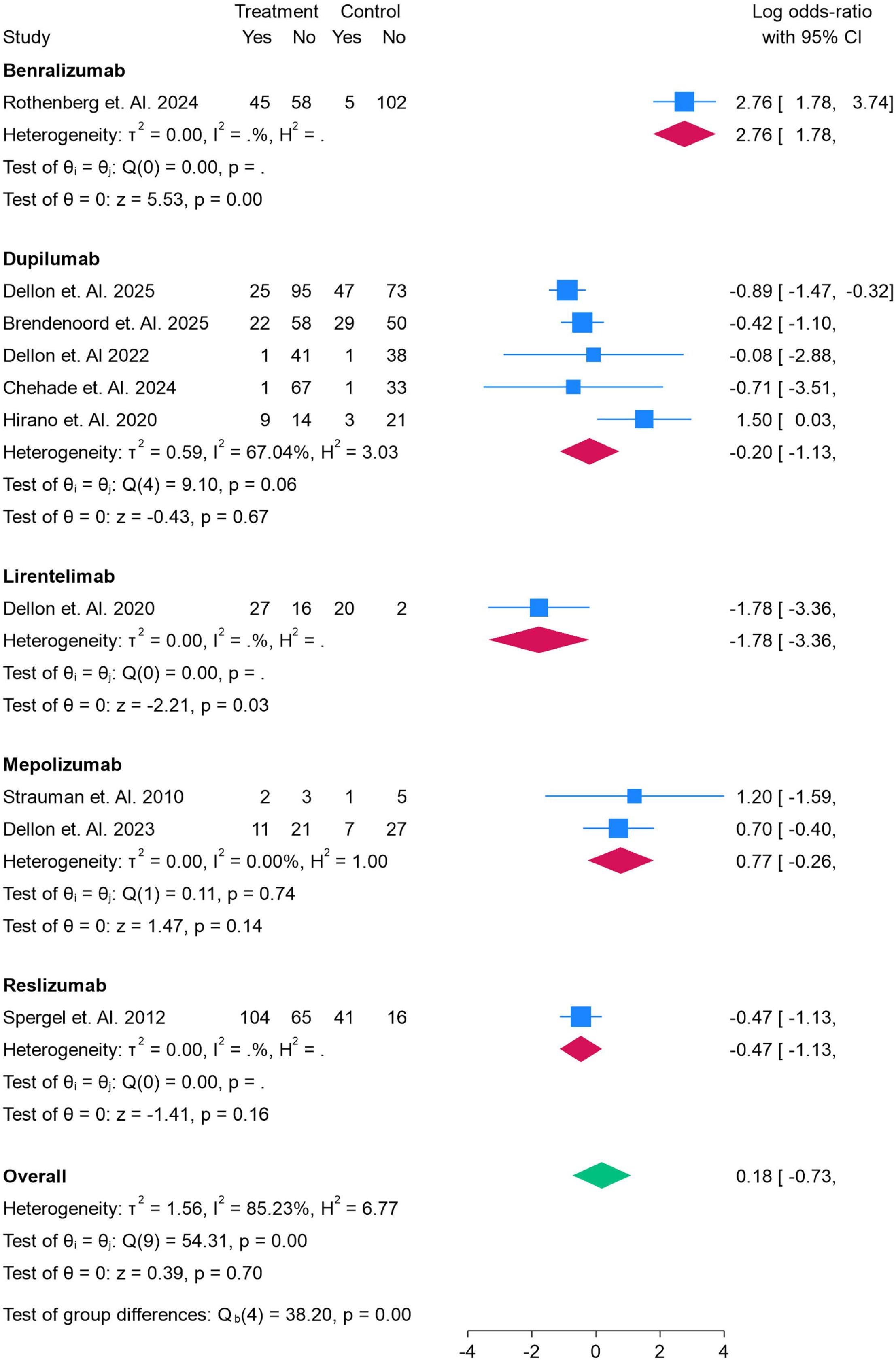
Symptomatic Response of the eosinophilic esophagitis in Patients

**Figure 4.**
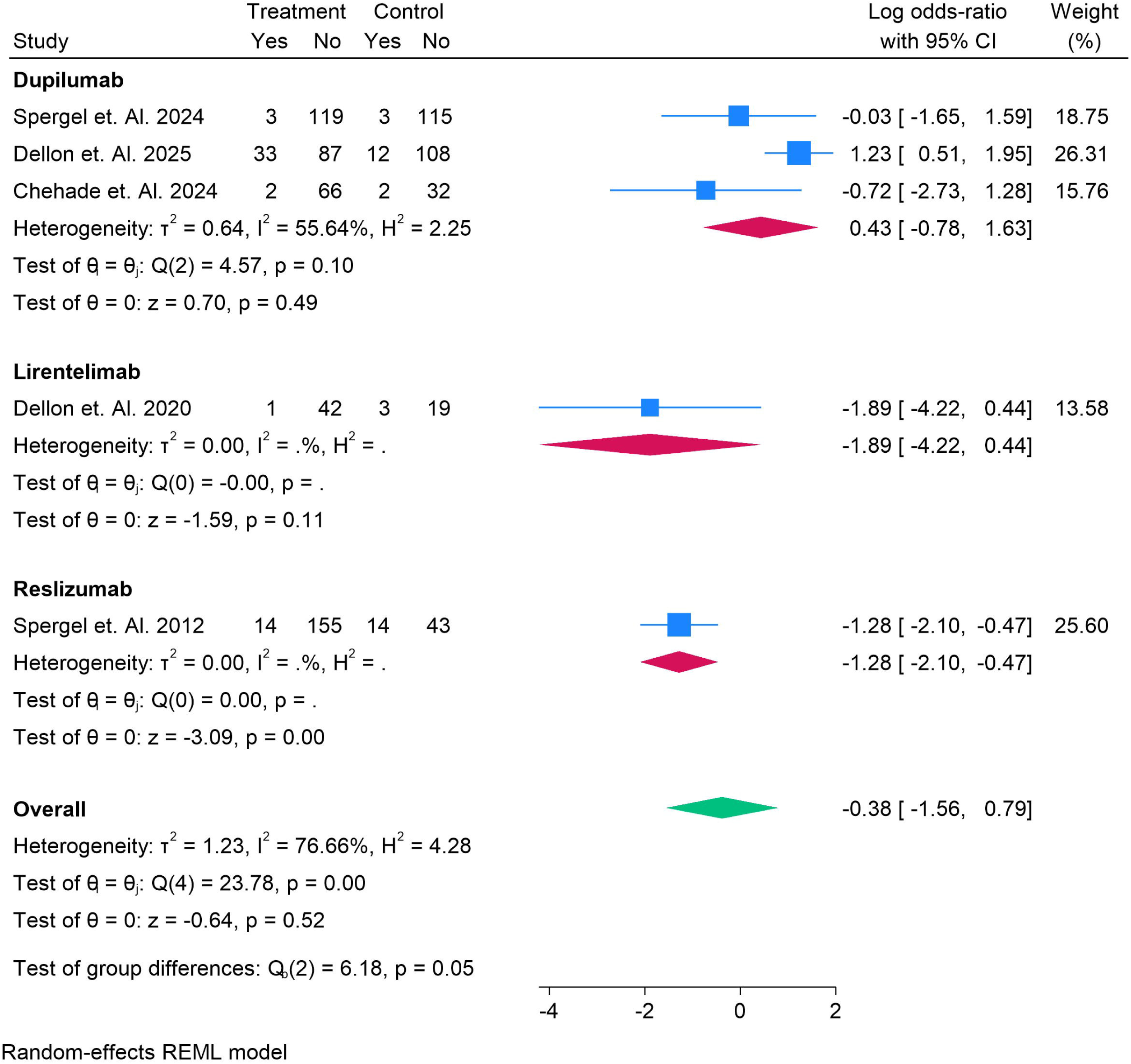
Endoscopic Remission of the eosinophilic esophagitis in Patients

**Figure 5.**
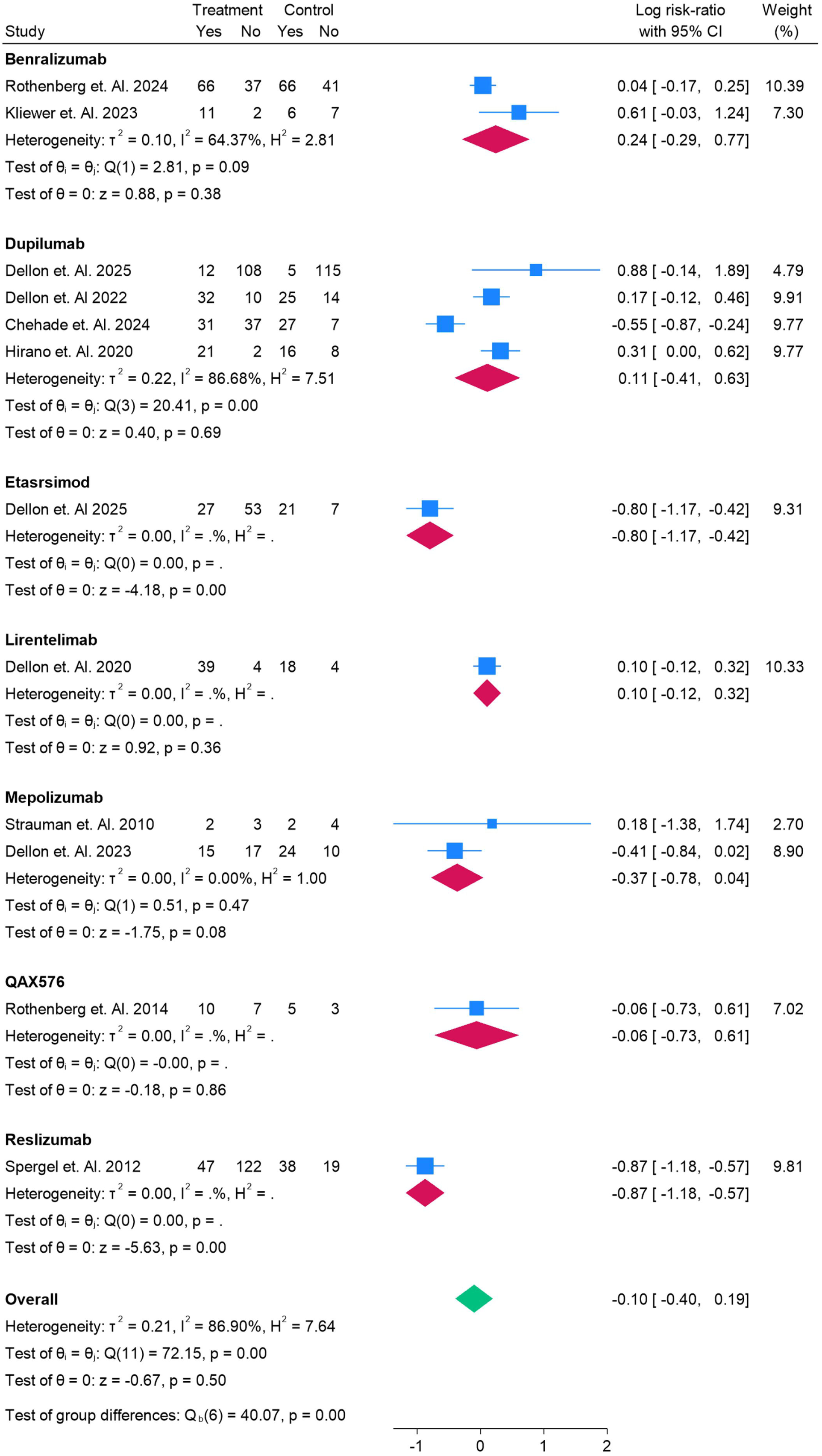
Any Adverse Events of the eosinophilic esophagitis in Patients

**Figure 6.**
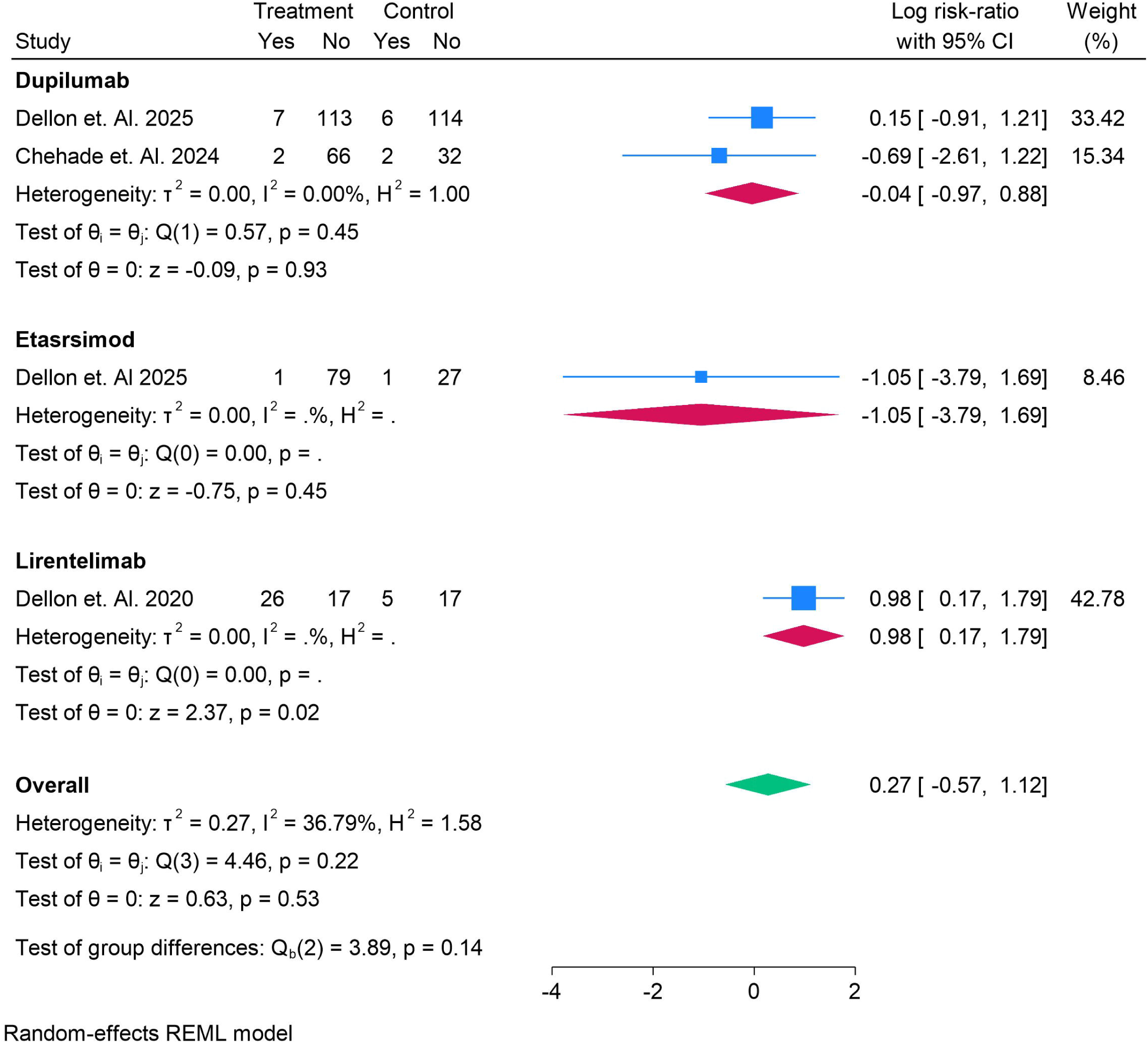
Cardiological Adverse Events of the eosinophilic esophagitis in Patients

**Figure 7.**
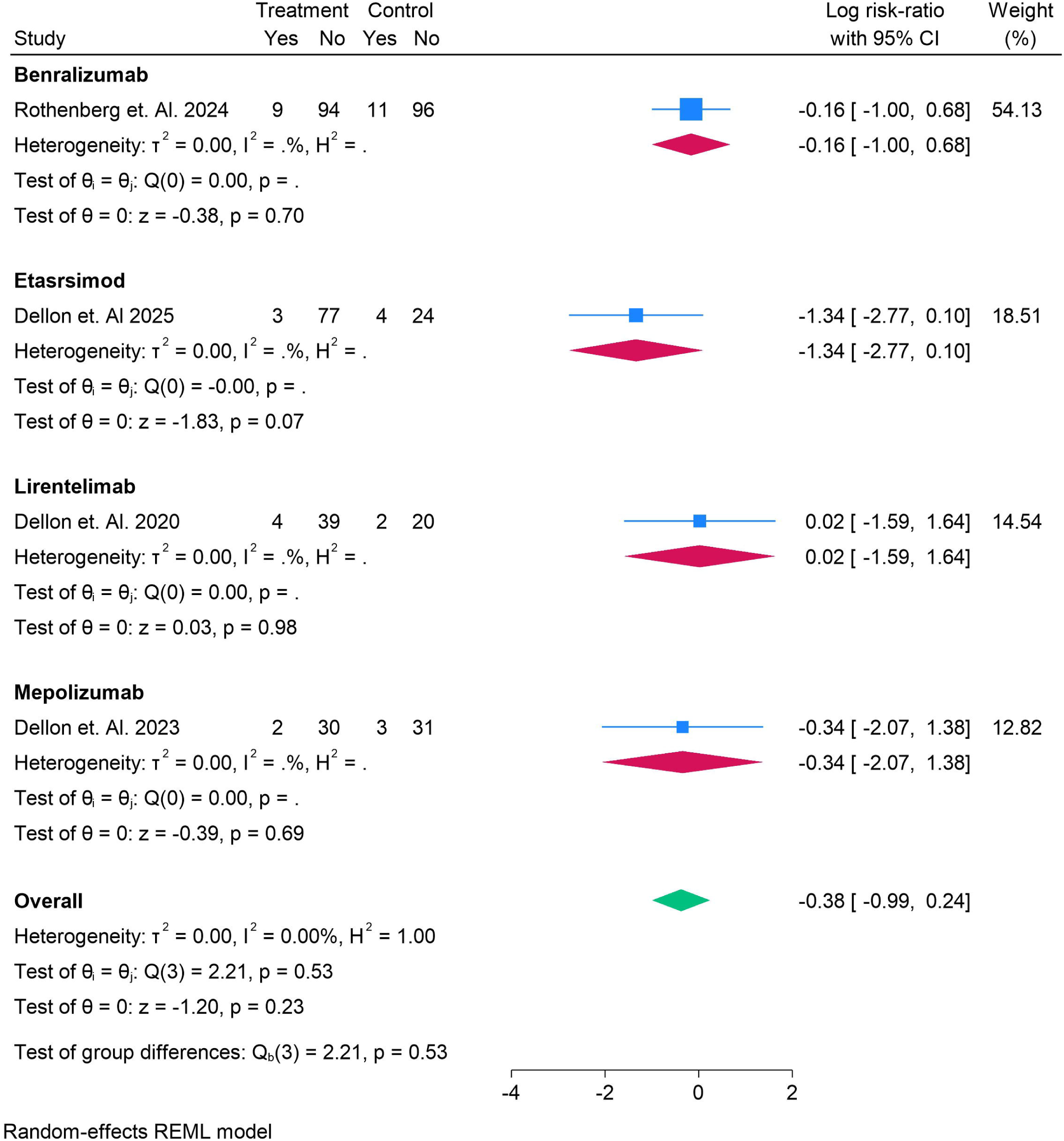
Neurological Adverse Events of the eosinophilic esophagitis in Patients

