## supplementary file for "Biologics for Eosinophilic Oesophagitis: A Systematic Review and Meta-Analysis of Randomized Controlled Trials"

Supplementary Files

Search String

Eosinophilic Esophagitis OR Eosinophilic Esophagitis OR EoE OR Eosinophilic Duodenitis OR Eosinophilic Gastritis OR Eosinophilic Gastroenteritis OR Eosinophilic Enteritis OR Eosinophilic Colitis OR Eosinophilic Gastrointestinal Diseases OR Eosinophilic Gastroenteropathy AND Biological Products OR Antibodies OR Biologics OR Cendakimab OR Dupilumab OR Mepolizumab OR Omalizumab OR Tezepelumab OR Bertilimumab OR Vedolizumab OR Tofacitinib OR Reslizumab OR IL-4 antibody OR IL-5 antibody OR IL-13 antibody OR IL-15 antibody OR IL-25 antibody OR IL-33 antibody OR eotaxin-1 antibody OR eotaxin-3 antibody OR CCR3 antibody OR Siglec-8 antibody OR IgE antibody OR TSLP antibody OR QAX576 OR RPC4046

Table S1. Summary Findings

| **Author Year** | **Country** | **Total Sample** | **Male** | **Female** | **Follow Up** | **Average Age** | **Diagnostic Criteria** | **Treatment Name** | **Treatment Number** | **Control Name** | **Control Number** | **Grade** | **Main Finding** |
| --- | --- | --- | --- | --- | --- | --- | --- | --- | --- | --- | --- | --- | --- |
| 12 Strauman et. Al. 2010 | Switzerland | 11 | 7 | 4 | 4.8 | 32.4 | PPI-refractory oesophageal symptoms **plus** oesophageal eosinophilia **>20 eosinophils/hpf** (with dysphagia). | Mepolizumab | 5 | Placebo | 6 | High | Mepolizumab **reduced oesophageal eosinophils and remodeling markers** but **did not achieve histologic remission or clear clinical benefit** versus placebo in this small adult EoO trial. |
| 13 Spergel et. Al. 2024 | USA | 240 | 120 | 120 | 6 | 35.6 | EoE diagnosed with peak eosinophils ≥15/HPF despite 8 weeks of high-dose PPI, and DSQ biweekly score ≥10 at baseline. | Dupilumab | 122 | Placebo | 118 | High | Dupilumab 300 mg weekly significantly improved HRQoL and symptoms—including beyond dysphagia—vs placebo by week 24 in adolescents/adults with EoE. |
| 14 Dellon et. Al. 2025 | USA | 240 | 120 | 120 | 6 | 33.5 | EoE described as chronic type-2 inflammatory disease with dysphagia and eosinophil-predominant esophageal inflammation; specific trial diagnostic inclusion criteria **not detailed** in this brief. | Dupilumab | 120 | Placebo | 120 | High | **Symptomatic improvement in EoE needs higher systemic dupilumab exposure than histologic response, supporting the approved 300 mg weekly regimen over q2w for symptom benefit.** |
| 15 Brendenoord et. Al. 2025 | USA | 159 | 80 | 79 | 6 | 31.2 | Adolescents ≥12 y and adults ≥18 y with documented EoE, PEC ≥ 15 eos/hpf after ≥8 weeks high-dose PPI; DSQ ≥ 10 at randomization. | Dupilumab | 80 | Placebo | 79 | High | Dupilumab 300 mg weekly improved histologic, symptomatic, and endoscopic outcomes vs placebo in EoE and remained effective through Week 52, irrespective of prior swallowed budesonide/fluticasone use, PPI use, elimination diet, or prior dilation. |
| 16 Dellon et. Al. 2020 | USA | 65 | 40 | 25 | 3.5 | 39 | Adults with moderate–severe GI symptoms plus mucosal eosinophilia: **≥30 eosinophils/HPF in ≥5 gastric HPFs and/or ≥30 eos/HPF in ≥3 duodenal HPFs**, with other causes excluded. | Lirentelimab | 43 | Placebo | 22 | High | AK002 (lirentelimab) **significantly reduced GI tissue eosinophils and improved symptoms vs placebo**, with more mild–moderate infusion-related reactions. |
| 17 Dellon et. Al 2022 | USA | 81 | 49 | 32 | 5.5 | 31.6 | EoE diagnosis: **esophageal mucosal biopsy with ≥15 eosinophils per high-power field with compatible clinical symptoms (no alternative causes)**. | Dupilumab | 42 | Placebo | 39 | High | Weekly dupilumab 300 mg **significantly increased histologic remission and improved dysphagia symptoms vs placebo at 24 weeks** (q2w improved histology but not symptoms per hierarchical testing). |
| 18 Chehade et. Al. 2024 | USA | 102 | 78 | 24 | 4 | 7.3 | Symptoms of esophageal dysfunction **and** esophageal mucosal biopsy with **≥15 eosinophils/high-power field** (no alternative causes); eligibility required ≥15 eos/hpf in ≥2 regions and **non-response after ≥8 weeks of PPI**. | Dupilumab | 68 | Placebo | 34 | High | Dupilumab induced **significantly higher histologic remission** than placebo at 16 weeks in children with EoE; the higher-exposure regimen also improved endoscopic and transcriptomic measures. |
| 19 Rothenberg et. Al. 2024 | USA | 211 | 157 | 53 | 6 | 33.7 | Symptomatic, histologically active EoE with ≥15 eosinophils per high-power field on centrally read biopsies at ≥2 esophageal levels, per consensus guidelines. | Benralizumab | 103 | Placebo | 107 | High | Benralizumab produced a very high histologic response versus placebo but did not improve dysphagia symptoms or endoscopic findings versus placebo at 24 weeks. |
| 20 Kliewer et. Al. 2023 | USA | 26 | 19 | 7 | 3 | 19.5 | Symptomatic, histologically-active EoG with **gastric eosinophil count ≥30 eos/hpf in ≥5 hpf** plus prior **blood eosinophilia >500/µL**. | Benralizumab | 13 | Placebo | 13 | High | **Benralizumab significantly increased histological remission vs placebo at 12 weeks (77% vs 8%) but did not improve symptoms or endoscopic/structural measures over the double-blind period.** |
| 21 Dellon et. Al. 2023 | USA | 66 | 39 | 27 | 6 | 33.1 | Age 16–75 with confirmed EoE per consensus guidelines, prior PPI-nonresponse, active eosinophilia with peak ≥15 eos/hpf, dysphagia (≥3 episodes over 2 weeks) and baseline EEsAI ≥ 27. | Mepolizumab | 32 | Placebo | 34 | High | Mepolizumab did **not** improve dysphagia symptoms vs placebo at 3 months, but **did** significantly reduce esophageal eosinophil counts and modestly improved endoscopic severity; extending to 6 months didn’t add benefit. |
| 22 Hirano et. Al. 2019 | Multicenter | 99 | 61 | 38 | 4 | 37.1 | Adults 18–65 y with dysphagia and **histologic EoE defined as peak ≥15 eosinophils/hpf at any 2 of 3 esophageal levels off anti-inflammatory therapy** (after a PPI trial). | RPC4046 | 65 | Placebo | 34 | High | **RPC4046 (anti-IL-13) significantly reduced histologic and endoscopic disease activity versus placebo after 16 weeks, with an overall safety profile similar to placebo.** |
| 23 Hirano et. Al. 2020 | Multicenter | 47 | 23 | 24 | 3.7 | 36.1 | Adults (18–65 y) with active EoE: **≥2 dysphagia episodes/week** and **peak esophageal eosinophil density ≥15 eos/HPF** on biopsies from **≥2 of 3 regions**, after **≥8 weeks of high-dose PPI**, per consensus guidelines. | Dupilumab | 23 | Placebo | 24 | High | Dupilumab **significantly improved dysphagia symptoms, histologic and endoscopic disease activity, and esophageal distensibility** versus placebo, with an **acceptable safety profile** in adults with active EoE. |
| 24 Spergel et. Al. 2012 | Multicenter | 226 | 172 | 54 | 3.5 | 12.7 | Symptomatic children/adolescents (moderate-or-worse symptom severity) with **≥24 intraepithelial eosinophils per high-power field** on esophageal biopsy and PPI trial ≥4 weeks without symptom resolution or normal pH probe. | Reslizumab | 169 | Placebo | 57 | High | Reslizumab significantly **reduced esophageal eosinophil counts** versus placebo, but **symptom improvements were similar across all groups** and did not correlate with histology. |
| 25 Dellon et. Al 2025 | Multicenter | 108 | 57 | 51 | 5.5 | 35.6 | Adults 18–65 y with prior EoE diagnosis and **histologically active disease (oesophageal peak eosinophil count ≥15 eos/hpf at screening EGD) plus DSQ dysphagia ≥2 episodes/week**, on stable diet/PPI as applicable | Etasrsimod | 80 | Placebo | 28 | High | **Etrasimod 2 mg** significantly reduced oesophageal peak eosinophil counts versus placebo by Week 16 (and further by Week 24), with acceptable tolerability. |
| 26 Rothenberg et. Al. 2014 | Multicenter | 25 | 24 | 1 | 6 | 30.3 | Adults (18–50 y) with PPI-resistant esophageal eosinophilia and peak eosinophil density ≥ 24 eos/hpf (400×) in proximal or distal esophagus (central pathology). | QAX576 | 17 | Placebo | 8 | High | QAX576 significantly reduced mean esophageal eosinophil counts and favorably shifted EoE-related gene expression with sustained effects, but the prespecified primary endpoint was not met; treatment was generally well tolerated. |

Risk of Bias

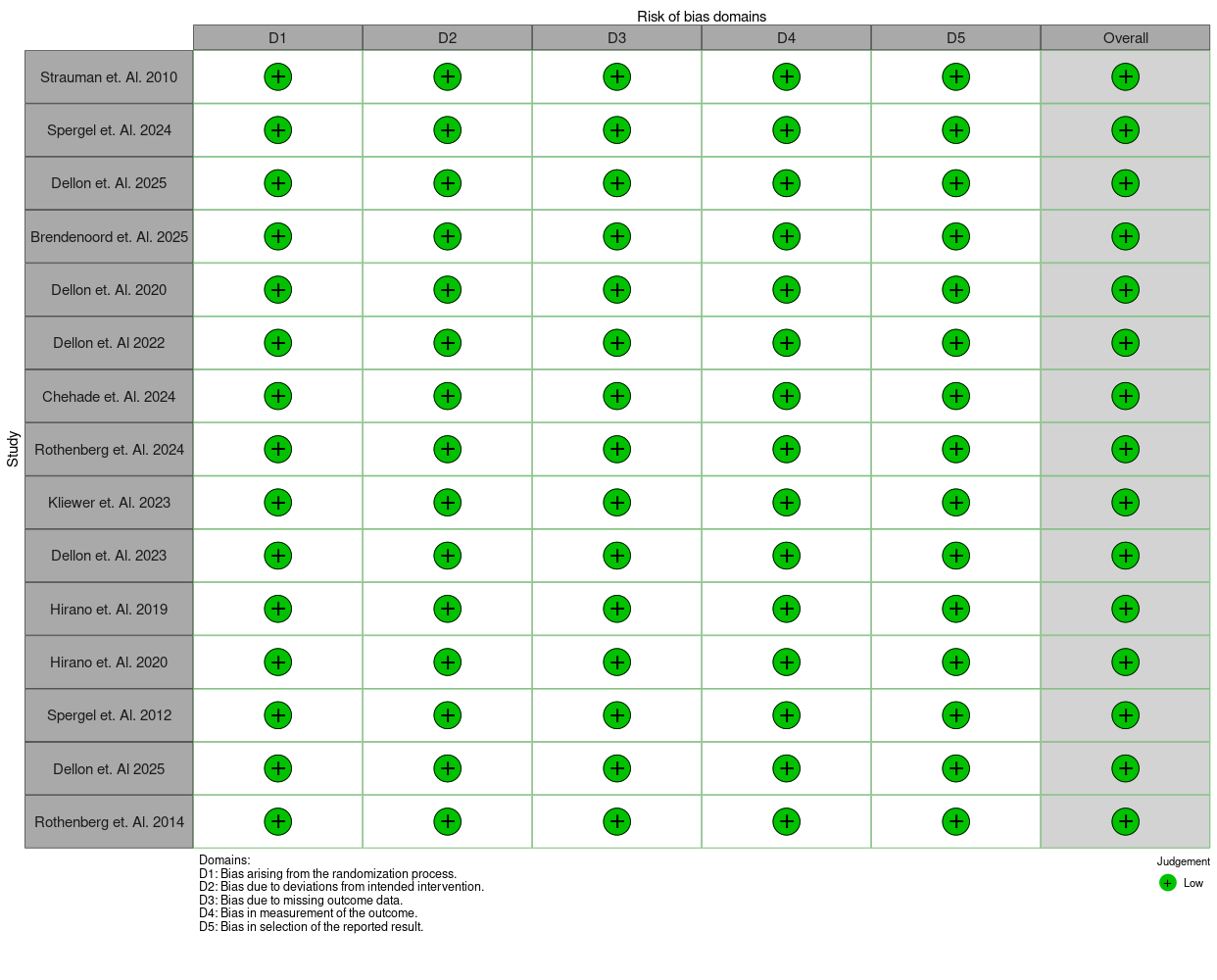
